# Quarantine and the risk of COVID-19 importation

**DOI:** 10.1101/2020.08.12.20173658

**Authors:** J. Arino, N. Bajeux, S. Portet, J. Watmough

## Abstract

Using a stochastic model, we assess the risk of importation-induced local transmission chains in locations seeing few or no local transmissions and evaluate the role of quarantine in the mitigation of this risk. We find that the rate of importations plays a critical role in determining the risk that case importations lead to local transmission chains, more so than local transmission characteristics, i.e., strength of social distancing measures (NPI). The latter influences the severity of the outbreaks when they do take place. Quarantine after arrival in a location is an efficacious way to reduce the rate of importations. Locations that see no or low level local transmission should ensure that the rate of importations remains low. A high level of compliance with post-arrival quarantine followed by testing achieves this objective with less of an impact than travel restrictions or bans.

## 1 Introduction

The spatio-temporal spread of COVID-19 is documented by a sequence of importation times reported at different administrative levels. See, for instance, the works [1-4] for accounts of early spread in several countries. Canada is used here as an example; the first confirmed case was reported in Ontario on 25 January 2020. British Columbia reported a case on 28 January 2020, but other provinces and territories (P/T) did not report cases until March or later, with the territory of Nunavut reporting its first confirmed case on 6 November 2020. Figure 1 shows the evolution of the percentage of jurisdictions reporting at least one new case in the past three weeks, for P/T and 112 Canadian health regions [5, 6].

**Figure 1:**
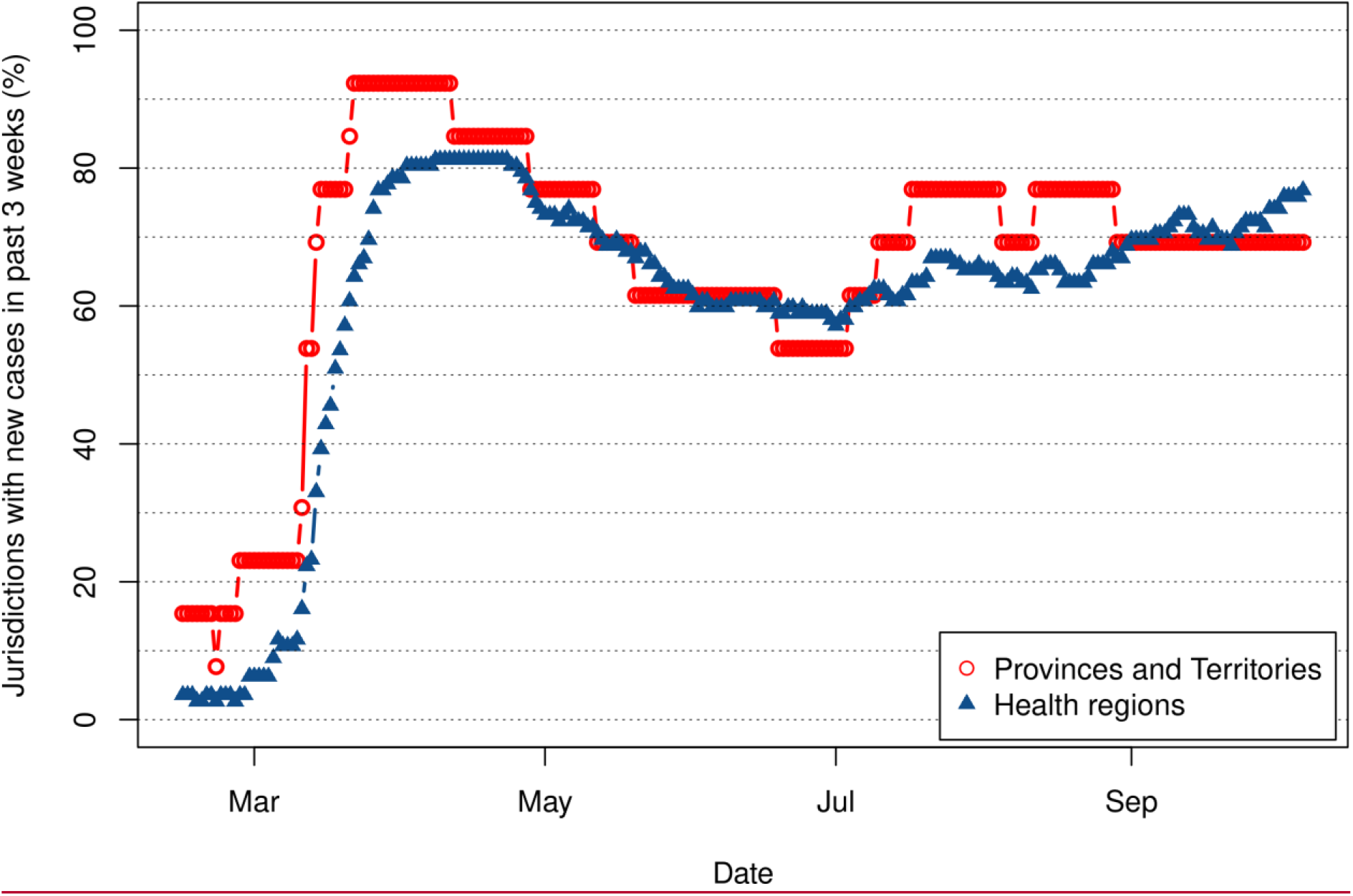
Percentage of provinces, territories and health regions in Canada having declared new confirmed cases of COVID-19 in 2020 during a three weeks period prior to dates shown. Time runs from three weeks after the first case in Canada on 25 January 2020 to 30 November 2020.

The initial increase in the number of jurisdictions affected was entirely driven by case importations to these jurisdictions. Later increases are indicative of importation of a case from another jurisdiction or existence of silent transmission chains lasting more than three weeks. We are concerned here with importations, whether in the initial stage or in later stages of the pandemic.

In the first phase of the pandemic, most countries (or lower level bodies in federal countries with devolved health care) took global, one-size-fits-all measures. After this initial lockdown period during which travel was also severely hampered, a second phase started, with local transmission down in some jurisdictions. However, with no therapeutic tools or vaccines yet available, transmission has picked up gain in some jurisdictions and many locations have been re-importing cases. It can be expected that until vaccines are widely deployed, many jurisdictions, in particular local ones, will experience such fluctuations. It seems important, in this context, to apply a more measured approach than the total top jurisdiction-level lockdowns that were used; the enormous economic cost of the first wave of lockdowns, combined with the potential diminishing compliance of individuals with public health measures, impose that public health authorities find ways to mitigate spread that are finer grained.

As part of the arsenal of measures available to public health authorities in the fight against the spread of COVID-19, there are some that are specifically geared towards the reduction of the risk of case importations: travel restrictions or bans, self-isolation upon arrival, etc. See the Electronic Supplement ES.1.1 for details. These measures have different effects and varying efficacy. In order to evaluate their relative effectiveness, it is important to better understand the importation process.

Some works [7-16] have considered the link between transportation and importation of COVID-19. We complement these works here by finely decomposing the process through which cases are imported into different types of events and focusing on the role of the rate at which a location is “challenged” by importations. We also consider the efficacy of the main method for reducing this rate while maintaining mobility: quarantine or self-isolation.

Importations are a critical component in the spatialisation of COVID-19 and other emerging or re-emerging infectious diseases, the overall spread phenomenon being driven by the repetition of *transport, importation, amplification* and *exportation* events. We further classify importations as *unsuccessful* if they do not lead to any local transmission chains or *successful* if they do. Successful importations then depend on the type of transmission chains they generate: they are *noncritical* or *critical* if they lead, respectively, to minor or major outbreaks. See Figure 2 and ES.1.1 for details.

**Figure 2:**
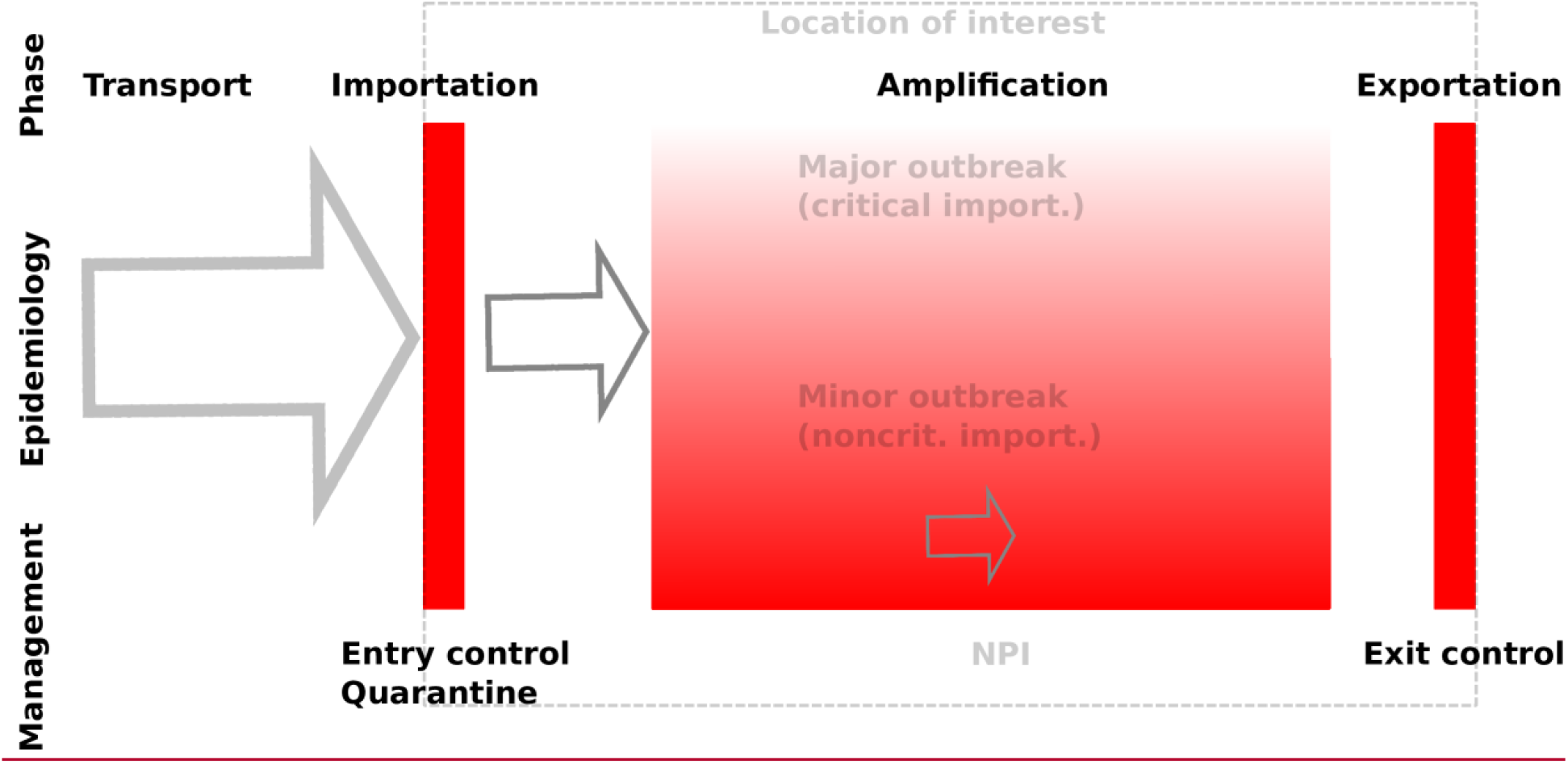
Spatialisation process: transport of cases from other locations; importation into the location of interest; local amplification; exportation to other locations. Red indicates control methods. See ES.1.1 for details.

Here, we consider importations of COVID-19 to locations that are not in the amplification stage and where importations could tip the balance in the direction of entering amplification. These include locations that have not had local cases yet, saw local transmission chains that have since extinguished or are seeing limited local transmission. The main issue tackled in this paper is the assessment of the risk that a successful importation occurs depending on the rate of case importations and local conditions, as well as the effect of post-arrival quarantine on these rates. To consider the problem, we use an SLIAR model “stimulated” by individuals flowing in from other locations.

## 2 Methods

We formulate the model as a continuous time Markov chain (CTMC) [17]. As the situation involves very small numbers of individuals, CTMC are preferable to ordinary differential equations, as they allow both integer counts of individuals and the incorporation of stochasticity. The setting under consideration is a single location in which the population is assumed to be homogeneously mixing. Initially, there are no active cases in the community; the model tracks the fate of the cohort of individuals who are susceptible to the disease. We consider the short term response of the model to stimulations taking the form of inflow of infected individuals, as schematised in Figure 3.

**Figure 3:**
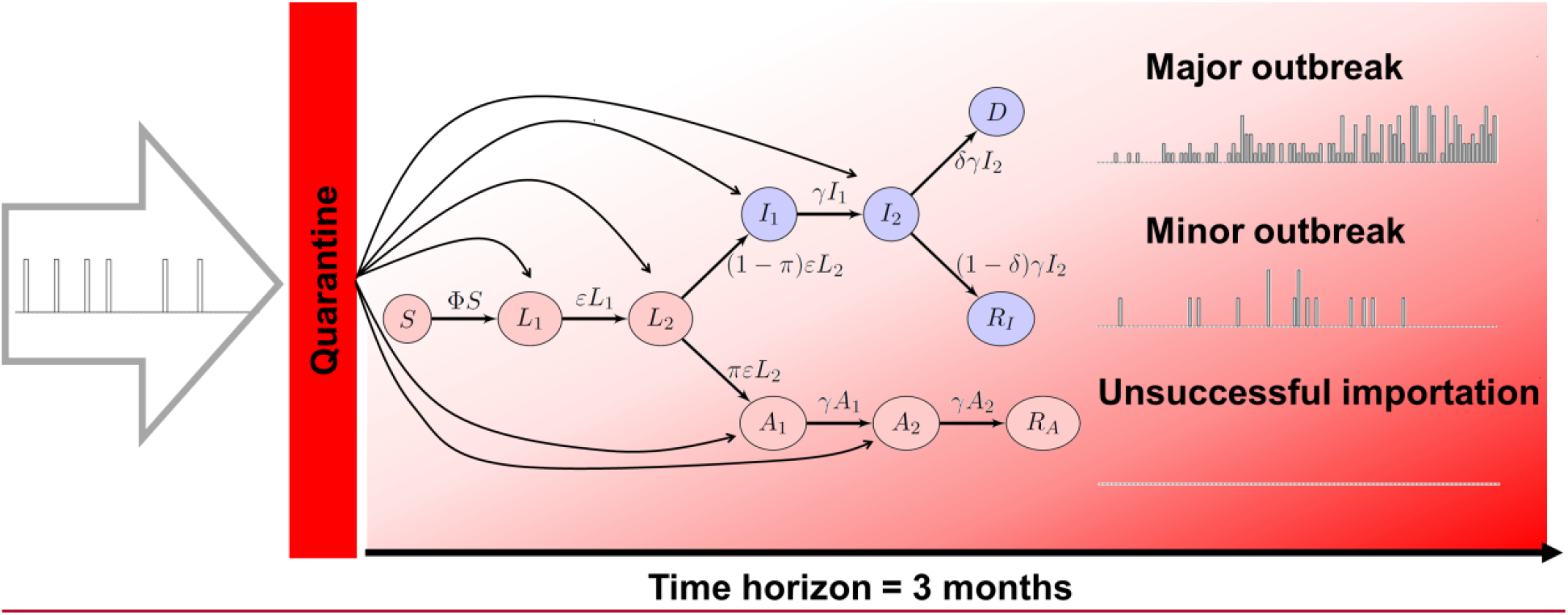
Input-output setting: imports arrive through transport, are potentially funnelled through quarantine, then enter one of the infected compartments. The three types of outcomes considered are shown on the right. In the epidemic model, compartment *S* represents suceptible individuals, *L*_1_ and *L*_2_ are latent individuals, *I*_1_ and I_2_ are detected infectious individuals, *A*_1_ and *A*_2_ are undetected infectious individuals, *D* are deaths from detected infections or posthumously tied to the infection; finally *R*_I_ and *R*_A_ are recovered from detected and undetected infections, respectively.

### 2.1 Epidemiological model and parameters

The structure of the model is detailed in ES.2 and ES.3, with the epidemiological model detailed in Figure ES.1: *susceptible* individuals, upon infection, move to the *latent* compartment. (Incubation and latent periods are assumed to overlap.) When their latent period is over, they can either progress to an *infection* that is ultimately *detected* or to one that remains *undetected*. At the end of the infectious period, individuals are *removed* (they *recover* or *die*); they no longer spread the disease. Post-recovery immunity is assumed to last at least as long as the (short) period of time under consideration.

We adopt a case detection-based approach. Detected infectious individuals are those who have been detected through testing, reporting or hospitalisation, i.e., individuals who appear as *confirmed* cases in the data. Undetected infectious individuals include those who are asymptomatic in the usual sense, but also symptomatic cases that avoid detection because of lack of testing. In the perspective of response to a crisis, using such a case detection-based definition allows to tailor model outputs to the situation in the location under consideration. This is further enhanced by using compartments for recovered individuals and death from detected infection directly connected to published data (Figures 3 and ES.1). One drawback from using this approach is that the some parameters (π, δ and η) incorporate not only disease characteristics but also some information about health policies specific to the location under consideration.

Simulations are tailored to locations, i.e., health regions or cities. The initial susceptible population, *S*(0), is the total population of the location under consideration, adjusted for pre-existing immunity if transmission occurred in the past. Epidemiological parameters are the means of those in [18] as well as values commonly found in the literature. Epidemiological and importation parameters are listed in Table 1. Simulations are run using the exact method in the R package *GillespieSSA2* with a time horizon of three months (92 days). Unless otherwise indicated, 1,000 simulations are run for each combination of parameter values and, when applicable, initial conditions.

**Table 1:**
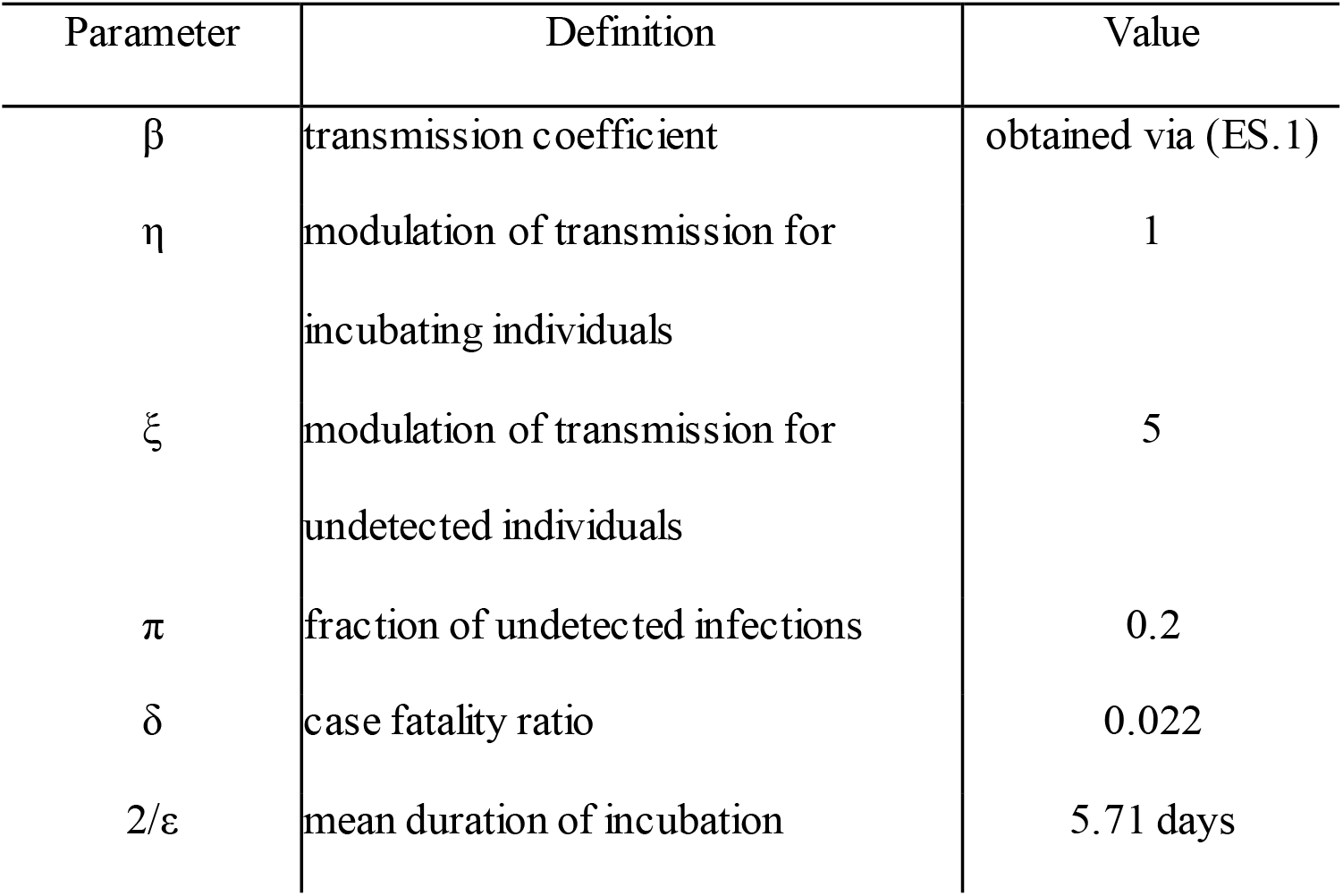
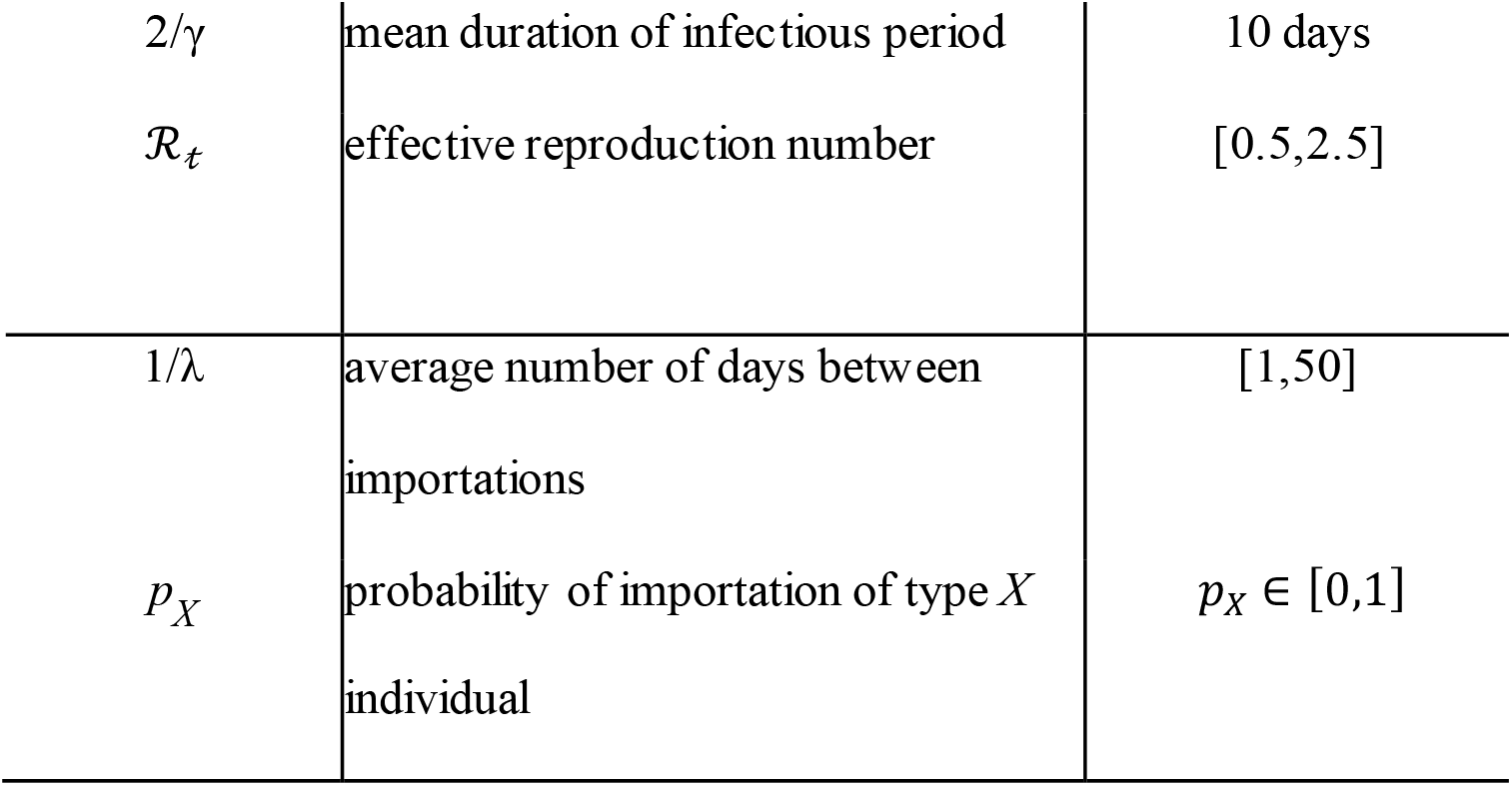
Model parameters.

| Parameter | Definition | Value |
| --- | --- | --- |
| $\beta$ | transmission coefficient | obtained via (ES.1) |
| $\eta$ | modulation of transmission for incubating individuals | 1 |
| $\xi$ | modulation of transmission for undetected individuals | 5 |
| $\pi$ | fraction of undetected infections | 0.2 |
| $\delta$ | case fatality ratio | 0.022 |
| $2/\epsilon$ | mean duration of incubation | 5.71 days |
| $2/\gamma$ | mean duration of infectious period | 10 days |
| $\mathcal{R}_t$ | effective reproduction number | [0.5,2.5] |
| $1/\lambda$ | average number of days between importations | [1,50] |
| $p_X$ | probability of importation of type $X$ individual | $p_X \in [0,1]$ |

### 2.2 Response of the system to case importations

Inputs to the system with transition probabilities (ES.3) are the *importation events*, i.e., stimulations taking the form of inflow of infected individuals as described by (ES.5); outputs are the responses of the system to these importation events. The nature of importation events and resulting outputs considered are summarised in Table 2.

**Table 2:** Simulation strategies. Questions addressed with the model and specification of inputs and outputs used. Individuals of type ℐ are those infected with the disease, i.e., ℐ ∈ {*L*_1_, *L*_2_, *I*_1_,*I*_2_, *A*_1_, *A*_2_}.

| Question | Input | Output |
| --- | --- | --- |
| SINGLE STIMULATION |  |  |
| Nature of importation | At $t=0$, importation of 1 individual of type $\mathcal{I}$<br>Model: (ES.3) | Probability of no outbreak, minor and major outbreak<br>First occurrence of $I_1$,<br>Time to local extinction, serial interval |
| Size of importation | At $t=0$, importation of $N$ individuals of type $\mathcal{U}$<br>Model: (ES.3) | Probability of no outbreak, minor and major outbreak |
| CHAIN OF STIMULATIONS |  |  |
| Rate of importation vs effective reproductive number | Importation of 1 individual of a type $\mathcal{U}$ with $p_{\mathcal{U}} = 1$ at rate $\lambda$<br>Model: (ES.3)+(ES.5) | Probability of no outbreak, minor and major outbreak<br>Attack rate |
| MODULATION OF CHAIN OF STIMULATIONS |  |  |
| Quarantine | $N$ individuals of type $\mathcal{U}$ arriving at rate $\lambda$ are quarantined for $t_q$ days<br>Model: (ES.3)+(ES.5) for $t_q$ days with $\beta=0$ | Quarantine efficacy<br>Quarantine-regulated importation rate |

Individuals of type 𝒰 are unobservable infected, i.e., 𝒰 ∈ {*L*_1_, *L*_2_, *A*_1_, *A*_2_}.

Of particular interest here is the consideration of these responses in terms of the severity of importations as defined in ES.1.1 and briefly summarised here.

- An *unsuccessful importation* has the import case not resulting in the transmission of the disease to anyone in the local population; in other words, there are no susceptible to latent (*S*→*L*_1_) transitions.
- A *successful importation* sees the import case resulting in at least one local case. Successful importations are further classified as follows.
  - A *noncritical successful importation* is one that is followed by a *minor outbreak*.
  - A *critical successful importation* is one followed by a *major outbreak*.

Minor and major outbreaks are defined as in a seminal paper of Whittle [19]; see details in ES.1.6. Finally, we say that the disease is *locally extinct* if there are no more cases of any type in the population under consideration.

## 3 Results

Table 2 summarises the questions investigated with the model and the outputs presented in the results.

### 3.1 Single stimulation simulations

#### 3.1.1 Role of the type of importation

The nature of the import case is important, as evidenced in Table ES.1. As can be expected, the earlier in the disease cycle an infected individual is when they are introduced in a population, the longer they spend interacting with others and thus the higher the risk that they transmit the disease. For instance, from Table ES.1, importing a single *A*_1_ undetected case is followed by a major outbreak in 27.5% (ℛ_*t*_ *=* 1.2) to 89.6% (ℛ_*t*_ *=* 2.5) of simulations, while importation of an *A*_2_ undetected case leads to a major outbreak in 17% (ℛ_*t*_ *=* 1.2) to 68.7% (ℛ_*t*_ *=* 2.5) of simulations. Earlier introduction in the latent stage lead to lower importation risks, because 80% of cases are detected and those cases are much less infectious (because detected cases are assumed to be mostly isolating). For comparison, a recent study by [15] found that a single importation leads to a large outbreak 17% to 25% of the time.

The nature of import cases is, to a large extent, a random hand that is dealt to importing jurisdictions. Unfortunately, most control measures they can implement “upstream” from an importation only have the capacity to remove symptomatically infectious individuals from the incoming flow, so that, in particular, latently infected individuals still can arrive.

#### 3.1.2 Role of the size of the importation

The size of the importation is naturally a key factor in the risk of importation, as was already established by [15]. To illustrate this and using outbreak severity criteria defined in ES.1.6, let us focus on importations of individuals in the *L*_1_ compartment. Qualitatively similar results are obtained by considering importations of other types of infected individuals but are not shown here.

Figure 4 shows the proportion of simulations with successful importations followed by major and minor outbreaks as a function of the importation size, i.e., the initial number of *L*_1_ individuals, for different values of the effective reproduction number ℛ_*t*_ chosen as illustrative of situations of lockdown (0.8), de-escalation of NPI measures (1.2) and uncontrolled spread (2.5). The proportion of simulations with critical successful importations, i.e., those followed by a major outbreak, is sensitive to the value of ℛ_*t*_. Therefore, the value of ℛ_*t*_ may significantly change the outcome if several infected individuals arrive simultaneously. The proportion of simulations with noncritical successful importations shows a maximum at a given initial number of *L*_1_. Indeed, when the initial number of *L*_1_ increases, it is more likely to trigger a critical successful importation than a noncritical one.

**Figure 4:**
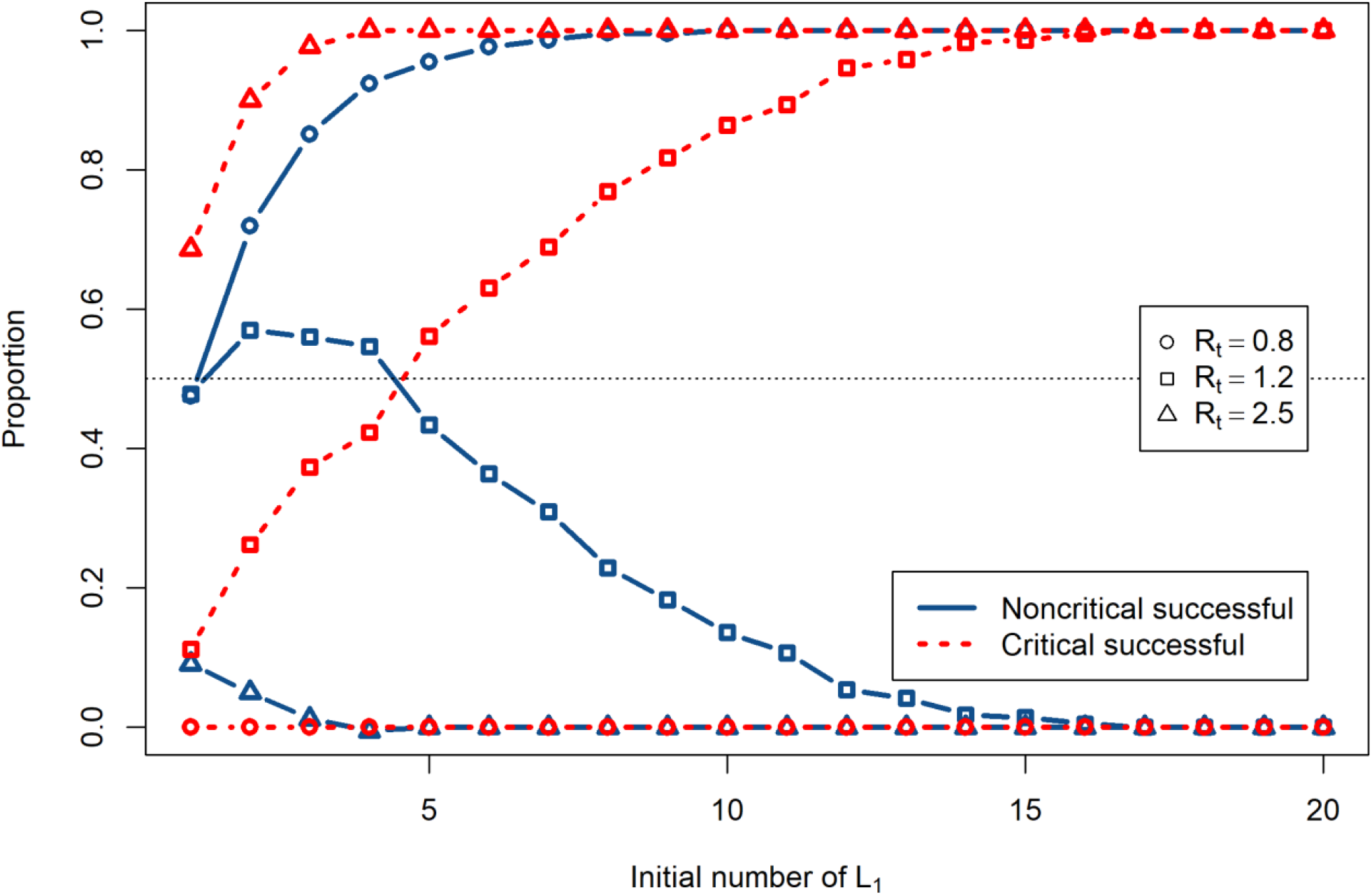
Proportions of critical (dotted red) and noncritical successful (solid blue) importations as a function of the initial number of imported *L*_1_ cases. Circles: ℛ_*t*_ *=* 0.8, squares: 1.2 and triangles: 2.5.

Here, there are obvious implications in terms of disease control, since one of the mechanisms leading to multiple simultaneous importations is infection during transport. Take for instance a location maintaining good but not perfect local conditions (ℛ_*t*_ *=* 1.2). If it receives four *L*_1_ individuals, then there is roughly a 50/50 chance that this leads to a major outbreak. If, on the other hand, these four individuals each infect another person during transport because of inadequate protocols aboard the incoming conveyance, then the odds of a major outbreak jump up to 3/4.

### 3.2 Effect of importation rates and NPI efficacy

We now consider the effect of repeated importations. We suppose that individuals arrive in the location through the importation process defined in (ES.5) by the Poisson distribution with parameter λ. The rate of importation λ from a given location can be approximated from epidemiological and travel characteristics of the origin location of the import case using (ES.6); the rate of importation from all sources is given in (ES.7). We vary 1/λ, the mean number of days between importation events. Here, 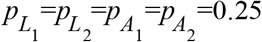 and 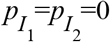 (we suppose detected infectious individuals are not able or allowed to travel).

Both the rate of case importations and the local effective reproduction number ℛ_*t*_ have an effect on the capacity of the disease to become established in the population. The larger ℛ_*t*_, the less efficacious the NPI. The raster in Figure 5 shows the proportion of simulations with a successful importation over a three month period.

**Figure 5:**
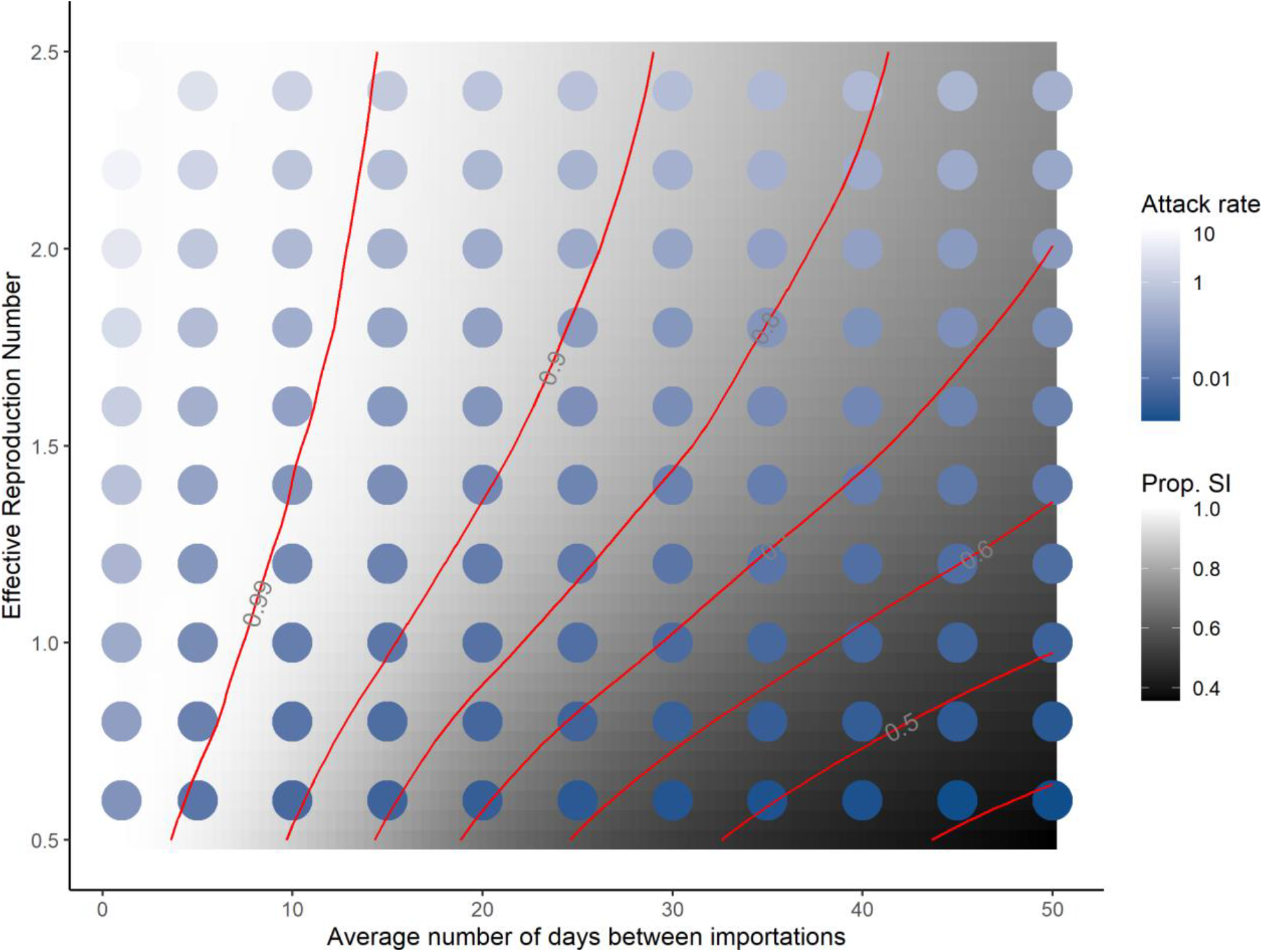
Proportion of successful importations (SI, grey scale) and attack rates (blue scale) in a 3 month period for different values of the average number of days 1/λ between two importations and ℛ_*t*_, an indicator of NPI efficacy.

When importations occur infrequently, local conditions are key. For instance, if importations occur on average every 40 days, local conditions change the risk of post-importation outbreaks over a three months period from roughly 40% to 80% of simulations. As the rate of case importations increases, post-importation outbreaks are increasingly likely for all local conditions, to the point that when cases are introduced every three days or less, 99% of simulations see outbreaks, regardless of the effort of local control. Thus, reducing the importation rate is key to preventing outbreaks.

On the other hand, the severity of the outbreaks is also sensitive to the value of ℛ_*t*_. The dots in Figure 5 show the attack rate of the disease in the population over the three months period considered, computed as the ratio (expressed as a percentage) (*S*(0) − *S*(*t*_*f*_)) /*S*(0), where *S*(0) and *S*(*t*_*f*_) are the number of susceptible individuals at the beginning and end of one simulation, respectively. Even though the probability of an outbreak is high when the rate of importations is high, regardless of local NPI effort, the intensity of local NPI effort greatly changes the outcome. Indeed, consider the attack rates where the average number of days between importations is one day (left-most column in Figure 5). Attack rates there range from 13.1% when ℛ_*t*_ *=* 2.5 to 0.02% when ℛ_*t*_ *=* 0.5. So, although the probability of importing the disease is roughly equal for this high importation rate, the severity of outcomes is very different. For ℛ_*t*_ > 1, we distinguish between minor and major outbreaks using the threshold τ defined in ES.1.6. This is shown in Figure ES.3.

### 3.3 Effect of post-arrival quarantine

The rate of importations plays a critical role in the risk that importations will trigger local transmission chains (Figure 5). The status of individuals when they arrive is also very important (Table ES.1). In order to evaluate the benefit of post-arrival quarantine, we proceed to the following simple numerical experiment. We consider Poisson generated chains of importation events, where each importation event is one of *L*_1_, *L*_2_, *A*_1_ or *A*_2_, i.e., one of the unobservable stages 𝒰. These chains are run through (ES.3) with no transmission (β=0) and for *t*_*q*_ days, where *t*_*q*_ is the duration of quarantine. Running the chains with no transmission means we consider the evolution of each individual case through disease stages during the quarantine period. Figure 6 shows, for quarantine periods of 7 and 14 days, the transitions between stages at the beginning and end of quarantine. We highlight in dark grey individuals who are still a risk to the community at the end of their quarantine period since they are still in unobservable stages 𝒰 (see ES.2.6 for details).

**Figure 6:**
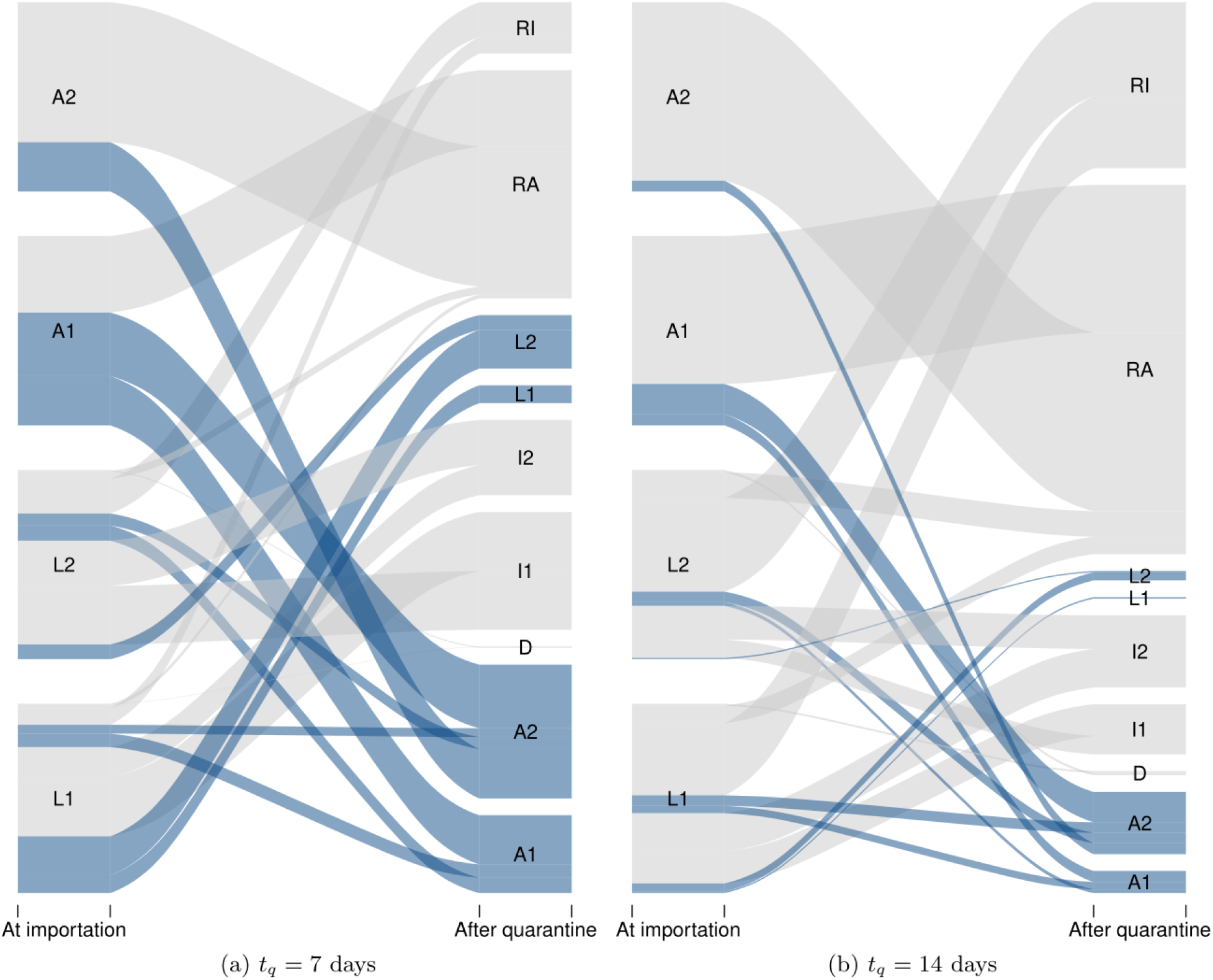
Evolution of the status of import cases during a (left) one or (right) two week quarantine period imposed upon arrival. Dark grey flows are individuals who are still a risk to the jurisdiction at the end of the quarantine period. Here, simulations were run for 2,500 individuals of each type of unobservable cases *L*_1_, *L*_2_, *A*_1_, and *A*_2_ entering quarantine.

In order to investigate the effect of the duration *t*_*q*_ of quarantine, as observed in Figure 6, we now quantify the efficacy of quarantine as the probability that a case that is initially unobservable becomes observable or recovers. Figure 7 shows the probability of success of quarantine (its efficacy *c*) as a function of its duration, for different values of the proportion π of undetected cases. The curves here are obtained by using the method in ES.2.6.

**Figure 7:**
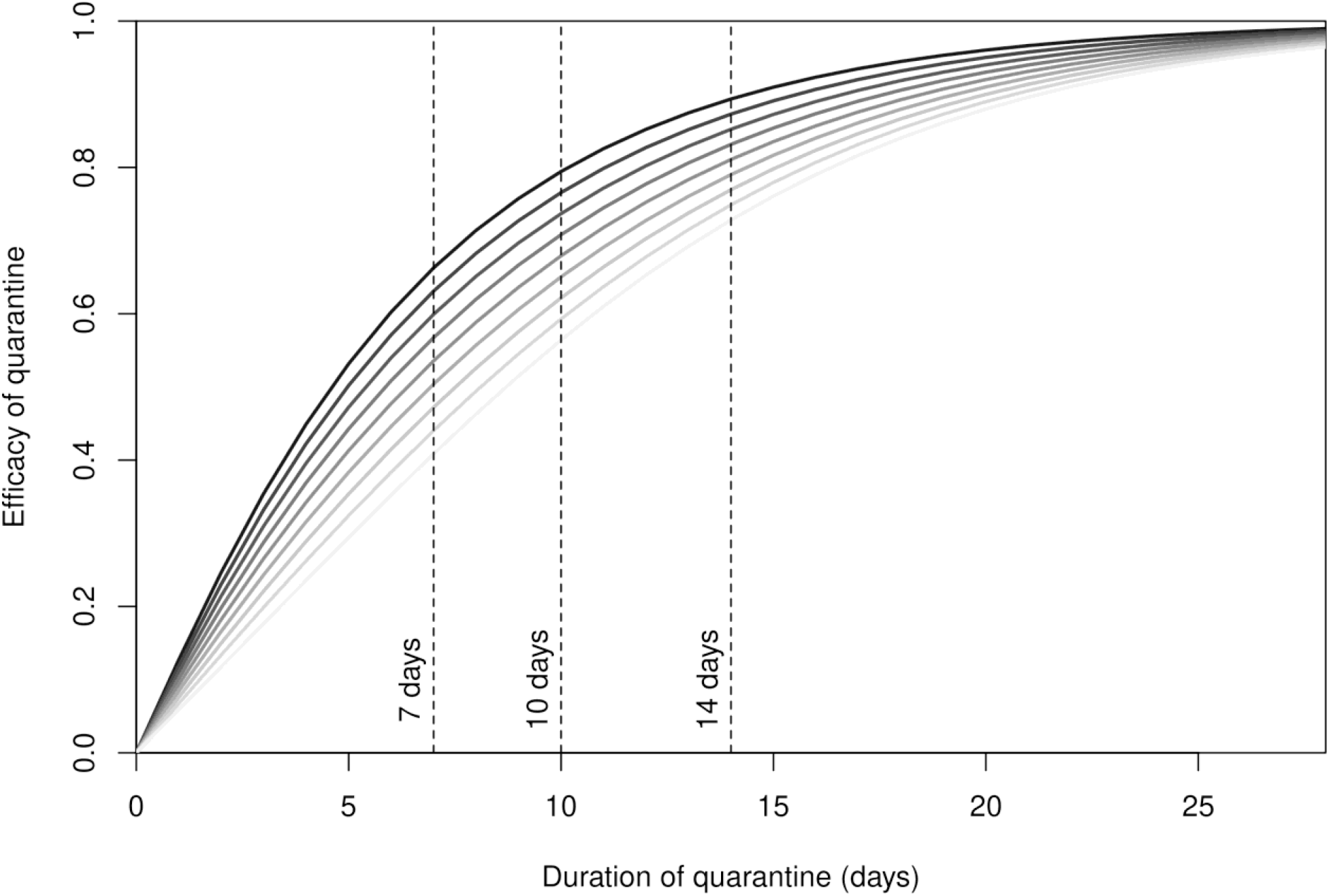
Probability *c* that the quarantine is successful as a function of its duration *t*_*q*_. From dark to light grey: fraction π of undetected cases varying from 0.1 to 0.9 by steps of 0.1. Vertical bars show the most commonly used quarantine durations.

We observe that the probability *c* of success of the quarantine increases with the duration of quarantine, as could be expected. Furthermore, from Figure 7, testing helps the success of quarantine. Indeed, consider for instance the most widely used duration of quarantine: two weeks. If 90% of cases are undetected, as could happen in a location making no effort to follow people during their isolation period, the efficacy of quarantine would be about 70%. Efficacy would reach 90%, on the other hand, if only 10% of cases went undetected.

Note that the effect of quarantine on the rate of importation is derived directly from the efficacy *c* of quarantine. If λ is the rate of importation prior to quarantine and λ_*q*_ is the quarantine-regulated rate of importation, then λ_*q*_=(1−*c*)λ. Consider Figure 5, whose abscissa is expressed in units of 1/λ. The effect of a quarantine with efficacy *c* is to scale the days between importations right by a factor of 1/(1−*c*). Consider a jurisdiction receiving a case on average every five days. A 50% efficacious quarantine leads to receiving one case every 10 days on average, while a 90% efficacious quarantine leads to receiving one case every 50 days on average.

## 4 Discussion

The main results of this study are highlighted by Figure 5, where the proportion of successful importations is given as a function of the effective reproduction number ℛ_*t*_ (used as a measure of the intensity of control efforts of local public health authorities) and the average number 1/λ of days between importations, and Figure 7, which shows the probability that quarantine is successful as a function of its duration *t*_*q*_.

The probability of an outbreak increases with the rate of importations, even when ℛ_*t*_ < 1, so that with importations every couple days or less, outbreaks are almost certain, regardless of local control efforts. However, the resulting attack rates increase with ℛ_*t*_, so local efforts always improve the outcome in that regard. For instance, with the parameters chosen for simulations, if case importations occur once every 10 days on average, then using measures to reduce ℛ_*t*_ to 0.5 still gives a 90% chance of outbreak (down from 99% when ℛ_*t*_ *=* 2.5), but with an attack rate of less than one percent at the end of three months (down from close to ten percent).

The sensitivity of the efficacy of quarantine to its duration and to monitoring effort seen in Figure 7 and the resulting scaling of importation rates have important policy implications. A location receiving few cases because it is connected only to places with zero or low prevalence could reasonably adopt a shorter (seven to ten day) quarantine period and still achieve a large right scaling on the importation risk (Figure 5), provided it also implements a high level of follow up of quarantining individuals (corresponding to lower values of π). Locations receiving cases at a high rate need to ensure that quarantine is longer and with high compliance with the imposed duration, since non-compliance is equivalent to imposing shorter quarantines.

Our results on importation risk apply to populations that see very few or no cases at all, or to places seeing more cases if appropriate contact tracing allows to distinguish between cases stemming from imported cases and community generated cases. Results on quarantine efficacy, on the other hand, apply in all circumstances since they do not involve transmission. Note that our results are also robust to the values used for parameters. We chose to use parameters mostly stemming from other work we have carried out on COVID-19 [18], but we could also have used values from the now abundant literature on the subject. This would modify some of the graphs in a limited manner but would not alter our conclusions.

Finally, note that the approach used here focuses on the rate at which cases are imported and is different from other published works on the topic, which amalgamate several distinct components. To get a sense of the rates one can expect to observe, a formula for determining the importation rates is provided in Section ES.1.5 and an example is given in Section ES.2.4, which allow to situate oneself within Figure 5. Also, one can use the calculator we provide in a Github repository to compute the efficacy of quarantine given by (ES.8) and the resulting quarantine-regulated importation rate (ES.9).

## Supporting information

Electronic appendix

## Data Availability

Some R code is available in a Github repository.

https://github.com/julien-arino/covid-19-importation-risk

## Data availability statement

Epidemiology and demography data used in this paper are publicly available on the Internet; sources are indicated in the references. The R code used in this paper is available on Github (https://github.com/julien-arino/covid-19-importation-risk) under an MIT license.

## Ethical approval

Not required.

## Contributors

All authors contributed equally to the work presented in this manuscript and to the preparation of this manuscript.

