## Supplementary material for "Quarantine and the risk of COVID-19 importation": Electronic appendix

J. Arino N. Bajeux S. Portet J. Watmough

### ES.1 Methods

#### ES.1.1 Mechanisms of spatialisation of epidemics and the nature of importations

We give here more details about the notions summarised in Figure 2 concerning the spatialisation of epidemics. Also, we make explicit the definitions regarding importations used in all the work.

*Transportation* is the actual movement of an infected individual to a new location. *Importation* occurs when an infected individual having acquired their infection in one location arrives in a different location while still infected with the disease. There can be two outcomes to an importation event; our terminology uses the point of view of SARS-CoV-2, the ætiological agent of COVID-19.

- An *unsuccessful* importation is one that does not lead to any further local transmission. Note that unsuccessful importations usually are not detected. It is possible that a location will see many unsuccessful importation events before it actually detects one.
- A *successful* importation is one that leads to at least one local transmission event, i.e., where the import case becomes the origin of one or more local transmission chains.

*Amplification* is then a critical phase of propagation that typically follows a successful importation. During amplification, cases multiply within a community, usually exponentially. In view of this, we define the *severity* of a successful importation as follows.

- A *noncritical successful importation* results in local chains of transmission that are not sustained, i.e., a *minor outbreak*. Case counts remain low before eventually dying out. As for unsuccessful importations, some of these local chains of transmission might go completely unnoticed.

- A *critical successful importation* sees sustained local chains of transmission. This is a *major outbreak*.

Finally, *exportation* is the process through which a community that is seeing some local transmission becomes the source of transportation events. Note that exportation does not require amplification to be taking place, although amplification does increase the probability that an individual leaving the community is harbouring the pathogen.

In our classification, exportation and transportation are very closely related. They are, however, considered as separate processes because they provide different entry points into the control of the spatial spread of COVID-19. Here, some methods of control of the spatial spread of an epidemic are listed.

- Acting on transportation involves limiting the number of individuals hailing from known exporting locations that are allowed to enter one's jurisdiction. In the case of COVID-19, this has taken the form of partial or complete travel bans.
- To minimize the chance that, were an importation to occur, it be a successful one, most jurisdictions imposed a quarantine for travellers inbound from other locations. Some jurisdictions also implemented entry screening.
- Once successful importations occur, jurisdictions used a variety of non-pharmaceutical interventions (NPI) to limit spread, i.e., curtail the amplification phase.

#### ES.1.2 Structure of the epidemiological model

The structure of the epidemiological model is a slight modification of the SLIAR model in [1], itself a modified version of the SLIAR model in [2]. However, we use a reinterpretation of the terms of the model to better correspond to the reality of COVID-19, although we keep the notation as in the papers above for simplicity. In short, rather than *symptomatic* and *asymptomatic* infections as in classical SLIAR models, we consider infections that are *detected* and *undetected*. In the perspective of response to a crisis, using a case detection-based definition allows to tailor model outputs to the situation in the location under consideration. We denote $I$ the detected individuals and $A$ the undetected ones. Detected infections are those that, at some point during the course of their infection or after death are identified through testing and are classified as *confirmed cases* in most of the data available online; the remainder of infections are undetected. Both detected and undetected infections can be symptomatic or asymptomatic cases. For instance, with low testing, some symptomatic cases might go undetected. Likewise, in the case of proper contact tracing, some asymptomatic cases may be detected.

The flow diagram is as shown in Figure ES.1: *susceptible* individuals, upon infection, move to the *latent* compartment. Incubation and latent periods are assumed to overlap. When their latent period is over, individuals can either progress to a detected infection or an undetected one. (Returning to a biological rather than observational definition of cases, it is generally assumed that in practice, there are more symptomatically infected individuals who are detected and more asymptomatic individuals who are undetected.)

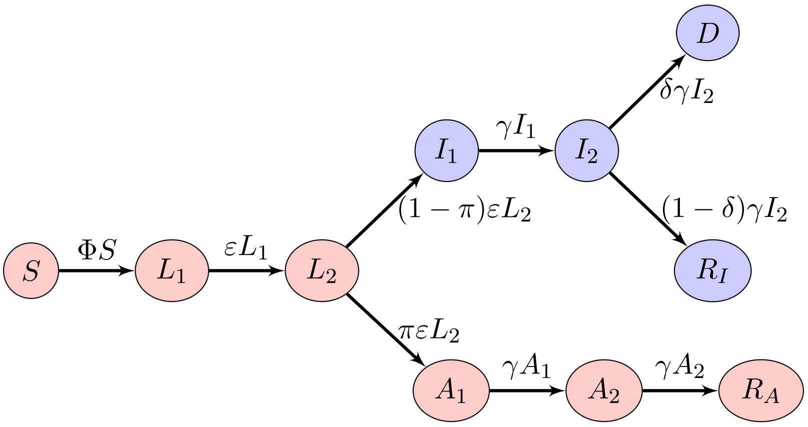


Figure ES.1: Flow diagram of the $SL_1L_2I_1I_2A_1A_2R_IR_AD$ model. $\Phi = \beta(I_1+I_2+\xi(A_1+A_2) + \eta L_2)$ is the force of infection. Blue compartments are observable (detected), with $I_1$ and $I_2$ usually indistinguishable and observed as $I_1+I_2$.

At the end of the infectious period, individuals are *removed*; they no longer spread the disease.

As in [1], there are two each of the latent, detected infectious and undetected infectious compartments. This is so that the time of sojourn in these disease states is Erlang distributed; see ES.2.3. Also, the force of infection $\Phi$ includes contributions not only from detected infectious $I$ and undetected infectious $A$ individuals but also from individuals in $L_2$. This is to accommodate observations of pre-symptomatic COVID-19 infections [3]. Finally, contrary to [1], the removed compartment is further subdivided into compartments for individuals having recovered from detected and undetected infections, $R_I$ and $R_A$, respectively, and those having died from the disease, $D$.

The case detection-based model we use here implies that some of the model parameters in [1] need to be reinterpreted. The mean duration of the incubation period, $2/\varepsilon$, is unchanged from the original SLIAR model; so is the mean duration $2/\gamma$ of the infectious period. The parameter $\delta \in [0,1]$ is the fatality ratio for detected cases, or, in other words, the case fatality ratio. Detection occurs for a fraction $1-\pi \in [0,1]$ of cases. Finally, $\eta$ and $\xi$ are modifiers of infectiousness due to being in late stage incubation and undetected, respectively.

Some remarks are in order concerning some of these parameters. First, concerning $\pi$, the fraction of cases undetected. Here, we think of detection as occurring throughout the course of the infection, not just at the end of the incubation period. This means that, in principle, there should be flows from the undetected compartments to the detected ones, e.g., from $A_1$ to $I_2$. However, our focus here is on the role of non-detection in the risk of importing COVID-19 cases, not on the specific role of testing. As a consequence, it is sufficient to make a phenomenological description of testing. A jurisdiction performing more testing will have lower values of $\pi$. Second, the parameters $\eta$ and $\xi$ play here a different role to the one they usually play in SLIAR models, because they modify the infectiousness of incubating or undetected individuals. In typical SLIAR models, they take values less than 1 because it is assumed that asymptomatically infectious individuals are less infectious than symptomatically infected ones. In the case of COVID-19, detected individuals typically have to self isolate or are hospitalised. As a consequence, these parameters can take here non-negative real values, not just values in $[0, 1]$.

Some notation is introduced to make discussions simpler. Individuals of type $\mathcal{I}$ are those *infected* with the disease, i.e., $\mathcal{I} \in \{L_1, L_2, I_1, I_2, A_1, A_2\}$. The prevalence of infection at time $t$ is then $\mathcal{I}(t) = L_1(t) + L_2(t) + I_1(t) + I_2(t) + A_1(t) + A_2(t)$. Individuals of type $\mathcal{U}$ are *unobservable* infected, i.e., $\mathcal{U} \in \{L_1, L_2, A_1, A_2\}$. *Observable* infected are $\mathcal{O} \in \{I_1, I_2\}$. The prevalence of unobservable and observable infections are obtained like that of infected cases, by summing the corresponding state variables. We can also define sets $\mathcal{U}_T$ and $\mathcal{O}_T$ for, respectively, total unobserved and observed cases, by adding $R_A$ to $\mathcal{U}$ and $R_I$ and $D$ to $\mathcal{O}$; these will not be used here.

We refer to [1] for some basic properties of the corresponding ordinary differential equations (ODE) model and just mention here that the effective reproduction number of the ODE version is

$$\mathcal{R}_t = \beta \left( 2\frac{\pi\xi}{\gamma} + 2\frac{1-\pi}{\gamma} + \frac{\eta}{\varepsilon} \right) S(0), \tag{ES.1}$$

where $S(0)$ is the initial susceptible population [1]. This formula is applicable to the stochastic model and is therefore useful to set some parameter values. It is used for instance when considering the intensity of NPI measures in the local community.

#### ES.1.3 Base model with a single importation event

Let $t \in [0, \infty)$ be a continuous variable (*time*). The random vector $\mathbf{V}(t)$ defined as

$$(S(t), L_1(t), L_2(t), I_1(t), I_2(t), A_1(t), A_2(t), R_I(t), R_A(t), D(t))$$

is the *state* of the system at time $t$. Denote $\Delta\mathbf{V}(t) = \mathbf{V}(t + \Delta t) - \mathbf{V}(t)$ the change in system state in the time interval $[t, t + \Delta t]$, with $\Delta t > 0$ sufficiently small to have at most one change during this interval. The probability of a transition is

$$\mathbb{P}\left(\Delta\mathbf{V}(t) | \mathbf{V}(t)\right), \tag{ES.2}$$

where $\Delta\mathbf{V}(t) = (\Delta S, \Delta L_1, \Delta L_2, \Delta I_1, \Delta I_2, \Delta A_1, \Delta A_2, \Delta R_I, \Delta R_A, \Delta D)$. The components $\Delta S, \Delta L_1, \ldots, \Delta D$ take only the values $\pm 1$ and 0 because of the hypothesis on $\Delta t$ being small enough. Transition probabilities are defined as follows (only nonzero values are shown):

| Rate | $\Delta S$ | $\Delta L_1$ | $\Delta L_2$ | $\Delta I_1$ | $\Delta I_2$ | $\Delta A_1$ | $\Delta A_2$ | $\Delta R_I$ | $\Delta R_A$ | $\Delta D$ |
|---|---|---|---|---|---|---|---|---|---|---|
| $\Phi S$ | $-1$ | 1 | | | | | | | | |
| $\varepsilon L_1$ | | $-1$ | 1 | | | | | | | |
| $(1-\pi)\varepsilon L_2$ | | | $-1$ | 1 | | | | | | |
| $\gamma I_1$ | | | | $-1$ | 1 | | | | | |
| $\delta\gamma I_2$ | | | | | $-1$ | | | | | 1 |
| $(1-\delta)\gamma I_2$ | | | | | $-1$ | | | 1 | | |
| $\pi\varepsilon L_2$ | | | $-1$ | | | 1 | | | | |
| $\gamma A_1$ | | | | | | $-1$ | 1 | | | |
| $\gamma A_2$ | | | | | | $-1$ | | | 1 | |

(ES.3)

Parameters are $\beta$, the transmission coefficient, $\eta$ and $\xi$ the modifications of infectiousness for individuals who are, respectively, pre-symptomatically and undetected infectious. A fraction $1-\pi$ of individuals are detected during the course of their infection or after dying from the disease (and correspondingly, $\pi$ are undetected).

Finally, $\varepsilon$ and $\gamma$ describe the rates at which incubation and infectiousness end, respectively. The fraction $\delta$ is the case fatality ratio.

Model (ES.3), when it is used to consider single introduction events, is combined with initial conditions at time $t=0$ of the form $S(0)=S_0$ (the initial susceptible population), $R_I(0)=R_A(0)=D(0)=0$ and one or several of $L_1(0), L_2(0), I_1(0), I_2(0), A_1(0)$ or $A_2(0)$ containing an integer number of individuals. In most cases, we consider the importation of a single individual, although in Section 3.1.2 we consider the effect of introductions of more than one infected individual.

When considering importations of a single infected individual at time $t=0$, explicit formula can be derived concerning probabilities of events affecting individuals. For instance, suppose that importation is of a single infected individual in the $L_2$ compartment, i.e., $L_2(0)=1$ with all other infected compartments empty. The probability that this individual recovers or dies from the disease without ever transmitting the virus is

$$\left(\frac{(1-\pi)\varepsilon}{\varepsilon+\beta\eta S(0)}\right)\left(\frac{\gamma}{\gamma+\beta S(0)}\right)^2+\left(\frac{\pi\varepsilon}{\varepsilon+\beta\eta S(0)}\right)\left(\frac{\gamma}{\gamma+\beta\xi S(0)}\right)^2. \tag{ES.4}$$

The first term is the probability of progression from $L_2$ to $I_1$ without local transmission. The second one is the probability of progression from $I_1$ to $D$ or $R_I$ without local transmission. The third term is the probability of progression from $L_2$ to $A_1$ without local transmission and the last term is the progression from $A_1$ to $R_A$ without local transmission.

#### ES.1.4 Adding repeated importations to the model

A case importation event is described by three components: the *time* at which the event occurs, the *size* of the importation (the number of cases imported simultaneously), and the *type* (epidemiological status) of cases that are imported. In order to better understand the role of the rate of importation and lower the number of parameters affecting the output of the model, we suppose here that all importations are of size 1 and focus on the rate (timing) and nature (type) of importations.

The timing of importations is described using a Poisson process with a parameter $\lambda$; the mean time between importation events is $1/\lambda$. An imported infected individual can belong to one of the six compartments $L_1$, $L_2$, $I_1$, $I_2$, $A_1$ or $A_2$; the infectious status of the imported individual is the type of the importation event. The probability of each type is given by $p_{L_1}$, $p_{L_2}$, $p_{I_1}$, $p_{I_2}$, $p_{A_1}$ and $p_{A_2}$ where $p_{L_1}+p_{L_2}+p_{I_1}+p_{I_2}+p_{A_1}+p_{A_2} = 1$.

So the model with repeated importations is the CTMC with transition rates given by (ES.3), with initial condition $S(0) = S_0$ and all other compartments zero, to which the following transitions are added.

| Rate | $\Delta L_1$ | $\Delta L_2$ | $\Delta I_1$ | $\Delta I_2$ | $\Delta A_1$ | $\Delta A_2$ |
|---|---|---|---|---|---|---|
| $\lambda p_{L_1}$ | 1 | | | | | |
| $\lambda p_{L_2}$ | | 1 | | | | |
| $\lambda p_{I_1}$ | | | 1 | | | |
| $\lambda p_{I_2}$ | | | | 1 | | |
| $\lambda p_{A_1}$ | | | | | 1 | |
| $\lambda p_{A_2}$ | | | | | | 1 |

(ES.5)

### ES.1.5 Estimation of importation rates

The number per day of case arrivals in *destination* from *origin* $i$ can be approximated by:

$$\begin{aligned} \lambda_i \simeq\ & \text{fraction active cases among population at } \textit{origin } i \\ \times\ & \text{fraction undetected among active cases at } \textit{origin } i \\ \times\ & \text{number of PAX per day from } \textit{origin } i \text{ to } \textit{destination}. \end{aligned} \tag{ES.6}$$

In general, a given location is connected to many other locations, say, $N$ of them. Using (ES.6), we obtain a value $\lambda_i$ for each of the $i = 1, \ldots, N$ locations that are potential sources of importation for the location under consideration. As arrival times are Poisson distributed, from the perspective of the receiving location, arrival times of events are independent exponentially distributed random variables. As a consequence, the parameter of the Poisson distribution for a location is obtained by considering competing risks and

$$\lambda = \sum_{i=1}^{N} \lambda_i. \tag{ES.7}$$

### ES.1.6 Characterising outbreak severity

To define the severity of the outcome, a threshold $\tau$ is chosen by adapting the results developed by Whittle [4] for a simpler stochastic SIR model. Whittle establishes that the probability of a *major* outbreak for an SIR model is $1 - (1/\mathcal{R}_t)^{I(0)}$ if $\mathcal{R}_t > 1$, while no major outbreak is possible when $\mathcal{R}_t < 1$, where $I(0)$ is the initial number of infected individuals in the population. He also establishes the probability of a minor outbreak when $\mathcal{R}_t < 1$, but this is not used here as we employ a different method for detecting minor outbreaks. Rather than computing the probability of a major outbreak, we decide on a probability $p$ (in practice, $p = 0.95$) of observing a major outbreak and set the threshold $\tau = -\ln(1-p)/\ln(\mathcal{R}_t)$. This threshold is then interpreted as follows: suppose that during a simulation, we observe a successful importation; if at some point following the importation, the prevalence of the disease increases to or past $\tau$, then with probability $p$, a major outbreak occurs.

As a consequence, we classify the type of importation events as follows.

- A successful importation sees an $S \to L_1$ transition for a local susceptible individual.
- An unsuccessful importation event does not lead to such a transition.
- A critical successful importation is detected by checking, when $\mathcal{R}_t > 1$, whether for some $t \in [0, t_f]$, the prevalence of infection $\mathcal{I}(t) > \tau$. There is no major outbreak (and thus no critical successful importations) for $\mathcal{R}_t < 1$.
- A noncritical successful importation is a successful importation that is not critical.

Severity can be further evaluated using observable infected cases $\mathcal{O}(t)$, unobservable infected cases $\mathcal{U}(t)$, and total prevalence of infection $\mathcal{I}(t)$.

### ES.2 Results

The examples that follow use the data for Prince Edward Island (PEI). Although not completely homogeneous, PEI is small in surface area (5,660 square kilometres) and quite densely populated (25 inhabitants per square kilometre). PEI can be reached by plane, ferry and through a bridge linking it to the continent (the Confederation Bridge). PEI has, to this point, had very few cases of COVID-19, most of them being imported.

#### ES.2.1 Role of the nature of the import case

Table ES.1 shows that the earlier in the course of the infection the imported case is, the more likely they are to lead to a successful importation and, conversely, importations in late stages of infection are less likely to be successful.

Note that the proportion of simulations with *unsuccessful importation* (UI) can be computed theoretically from equations such as (ES.4). These values can be compared to those in Table ES.1, which are obtained by simulation, giving a good agreement (not shown). In further simulations, we assume that $I$ individuals cannot travel. Note that we take sample values of $\mathcal{R}_t$ somewhat arbitrarily, reasoning as follows. The value $\mathcal{R}_t = 2.5$ was a value that was often given during the early days of the pandemic. So for us, this represents a non-contained epidemic where public health authorities are doing little to curtail spread. This is also the maximum value for the range of $\mathcal{R}_t$ we consider. If public health authorities take measures but those measures are not entirely satisfactory, then

| | $\mathcal{R}_t$ | Compartment with initial case $L_1$ | $L_2$ | $A_1$ | $A_2$ |
|---|---|---|---|---|---|
| UI | 0.8 | 0.56 | 0.571 | 0.248 | 0.489 |
| – | 1.2 | 0.455 | 0.438 | 0.173 | 0.402 |
| – | 2.5 | 0.235 | 0.245 | 0.062 | 0.234 |
| NSI | 0.8 | 0.44 | 0.429 | 0.752 | 0.511 |
| – | 1.2 | 0.429 | 0.440 | 0.552 | 0.428 |
| – | 2.5 | 0.103 | 0.113 | 0.042 | 0.079 |
| CSI | 0.8 | 0 | 0 | 0 | 0 |
| – | 1.2 | 0.116 | 0.122 | 0.275 | 0.170 |
| – | 2.5 | 0.662 | 0.642 | 0.896 | 0.687 |

Table ES.1: Proportion of simulations with unsuccessful (UI), noncritical (NSI) and critical (CSI) successful importations, as a function of the type of initial case.

the epidemic would still spread, albeit at a much lower rate. This is the value $\mathcal{R}_t = 1.2$. Finally, if efforts are satisfactory but not perfect, then the epidemic tends to die out, so $\mathcal{R}_t = 0.8$.

#### ES.2.2 Time to detection of local cases after importation

We continue with single stimulation simulations and present here results not discussed in the main text. We take the example of importation at time $t = 0$ of an $L_1$ individual; results are qualitatively similar for importations of other types, with only the time distributions varying.

In Figure ES.2, the first violin plot describes the distribution of local extinction times, the second shows the time distribution of the first *local* detected case, the third represents the time distribution of the first *import* detected case, i.e. when the import case $L_1$ becomes $I_1$. The distribution of serial intervals generated by the index case is shown on the right. Here, we define the serial interval as the length of time between the start of the simulation (i.e., the importation of an $L_1$ individual) and the time at which the first local infection occurs ($S \to L_1$ transition).

The median time to the detection ($I_1$ individual) following an importation event is 4.7 days (95% confidence interval $1.01 - 13.6$) for the import case and 15.3 days (95% confidence interval $6.04 - 30.6$) for a local case. In contrast, the first local infection event ($S \to L_1$ transition) occurs, on average, 9.4 days following an importation event (95% confidence interval $2.7 - 21.8$), and more than 81% of these first infections occur after 5 days (roughly the median time of the first $L_2 \to I_1$ transition).

Thus, if the import case is detected, this happens generally slightly before the first local transmission event takes place. In such a situation, the immediate isolation of the infected import case can prevent over half the local transmissions this individual would make. Note that this highlights the importance of quarantine: those transmissions that happen before isolation would not have taken place had this individual been quarantined. So quarantine is an efficacious way to compensate for delays in detection and isolation.

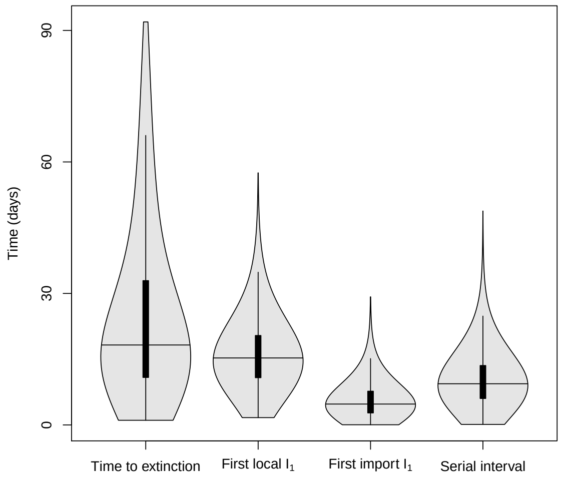


Figure ES.2: Time to extinction of the disease, first detected locally infected individual, first detected imported case, and serial interval. Base case simulation with $S(0) = 142,906$ (population of Prince Edward Island), $L_1(0) = 1$ and all other states equal to 0. Horizontal lines indicate the median value. Black rectangles are the inter-quartile range. Here, $\mathcal{R}_t = 0.8$.

Most first detections of local $I_1$ take place within 20 days following the importation of an $L_1$ individual; however, in some situations, the first infected and detected $I_1$ individual appears more than 30 days after the importation event. This characterises silent local transmission chains, i.e., ones involving only undetected infected individuals.

#### ES.2.3 Effect of the distribution of incubation periods

As mentioned in Section 2.1, compartments for $L$, $I$ and $A$ are subdivided in order to have Erlang distributed times of sojourn in these compartments rather than exponentially distributed ones. For instance, an individual traversing the two compartments $L_1$ and $L_2$ at the rate $\varepsilon$ spends an average time $2/\varepsilon$ between entry into $L_1$ and exit from $L_2$, with their time of sojourn Erlang distributed.

Between the incubation and infectious periods, the average COVID-19 patient spends an average of 15 days infected. The time horizon for the present work and for other work on COVID-19, on the other hand, is short. With such commensurate time scales, the variance of distributions becomes important. Several papers have considered distributions of incubation periods [5, 6]. Early work on 181 patients outside Hubei province before 24 February 2020 [5] found the best fit for the distribution of incubation times for these patients to be an Erlang distribution with shape parameter (the number of compartments needed in our approach) equal to six. In order to judge the effect of using more compartments to obtain a better shaped distribution of incubation times, we ran the same simulations as in Table ES.1, with six $L$ compartments instead of two as in the rest of this work. Results are shown in Table ES.2; the model is robust to the number of latent compartments used.

| | | Compartment with initial case | | | |
|---|---|---|---|---|---|
| | $n$ | $L_1$ | $L_2$ | $A_1$ | $A_2$ |
| UI | 2 | 0.401 | 0.434 | 0.757 | 0.848 |
| – | 6 | 0.436 | 0.392 | 0.746 | 0.836 |
| NSI | 2 | 0.484 | 0.458 | 0.208 | 0.132 |
| – | 6 | 0.464 | 0.513 | 0.225 | 0.151 |

Table ES.2: Proportions of simulations with unsuccessful (UI) and noncritical successful (NSI) importations, as a function of the type of initial case, when $R_t = 1.2$. $n$ is the number of latent compartments used in the simulations.

#### ES.2.4 Estimation of importation rates

As some of our numerics is inspired by the example of Prince Edward Island (PEI), let us continue with this. The first day of the "Atlantic bubble", where residents of all Maritime provinces of Canada were allowed to travel freely between these provinces, 8,500 people used the Confederation Bridge, which links PEI to the continent (in New Brunswick), going towards PEI. Suppose that cases in this flow happen with the same rate as in the general population and that prevalence in the general population can be deduced from confirmed case counts.

Assume that all travellers to PEI that day came from New Brunswick and that this relaxation of travel restrictions happened when New Brunswick experienced its highest number of active cases, on 2 April 2020, when there were 72 active cases [7]. The estimate of the population of New Brunswick for the second quarter of 2020 was 780,890 [8], i.e., a fraction of active cases of 0.000092202. Estimates for the prevalence of asymptomaticity and non-detection vary widely, with the latter being to a large extent driven by testing effort. Let us use a figure that seems reasonable at that stage of the pandemic, when New Brunswick had very few cases and where asymptomaticity was therefore the main driver for non-detection: 20%. (At the time of writing, New Brunswick still has very few cases.) Based on (ES.6), this gives 0.156744228 expected cases that day or, in other words, a mean time between importations of about 6.4 days, implying a rather high risk of importation (see Figure 5).

In practice, the Atlantic Bubble was put in place on 3 July 2020, at which time there was 1 active case in New Brunswick. Reasoning the same way, the mean time between importations in this instance would be of about 460 days, giving a very low risk of importation.

#### ES.2.5 Effect of importation rates on outbreak severity

Using the threshold $\tau$ derived from Whittle [4], we investigate in Figure ES.3 the proportion of simulations with critical successful importations when $\mathcal{R}_t > 1$.

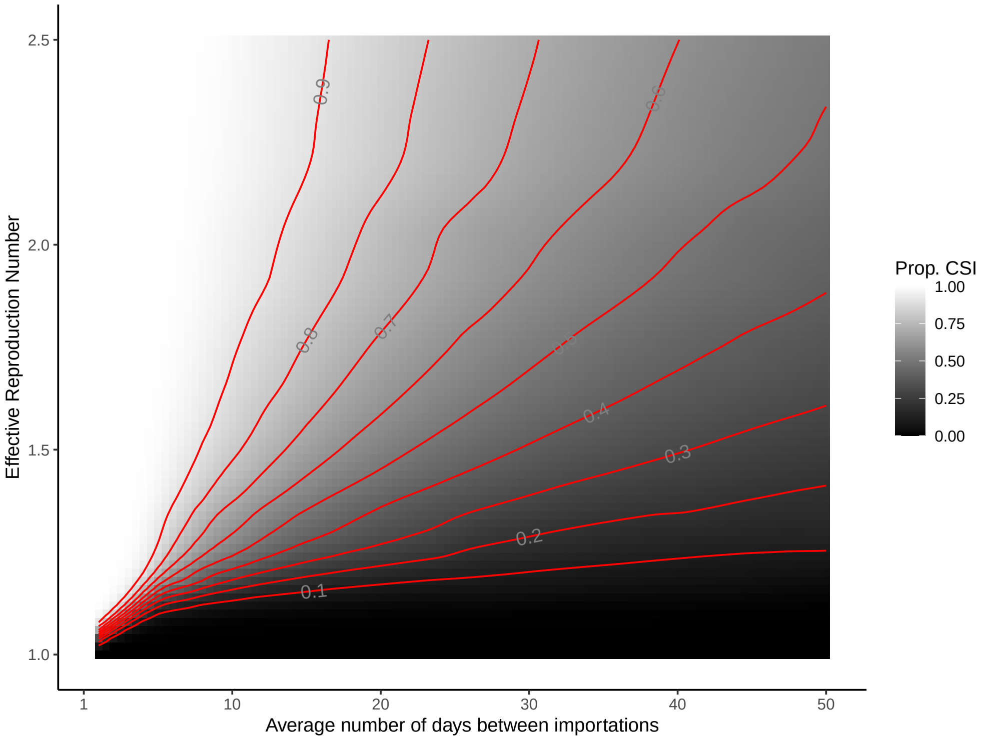


Figure ES.3: Proportion of critical successful importations (CSI) in a 3 month period for different values of the average number of days between two importations and $\mathcal{R}_t$, an indicator of NPI efficacy.

#### ES.2.6 Effect of post-arrival quarantine

The efficacy of quarantine can be expressed as the complement of the probability that an imported case is still in one of the unobservable infected states $L_1$, $L_2$, $A_1$, or $A_2$ at the end of the quarantine time $t_q$, i.e., the probability they emerge from quarantine infected and undetected. Consider the matrix of transition rates constructed from the entries of the first column of (ES.3), represented here as the table of transition rates between stages of infection:

| | $L_1$ | $L_2$ | $I_1$ | $I_2$ | $A_1$ | $A_2$ | $R_I$ | $R_A$ | $D$ |
|---|---|---|---|---|---|---|---|---|---|
| $L_1$ | $-\varepsilon$ | 0 | 0 | 0 | 0 | 0 | 0 | 0 | 0 |
| $L_2$ | $\varepsilon$ | $-\varepsilon$ | 0 | 0 | 0 | 0 | 0 | 0 | 0 |
| $I_1$ | 0 | $(1-\pi)\varepsilon$ | $-\gamma$ | 0 | 0 | 0 | 0 | 0 | 0 |
| $I_2$ | 0 | 0 | $\gamma$ | $-\gamma$ | 0 | 0 | 0 | 0 | 0 |
| $A_1$ | 0 | $\pi\varepsilon$ | 0 | 0 | $-\gamma$ | 0 | 0 | 0 | 0 |
| $A_2$ | 0 | 0 | 0 | 0 | $\gamma$ | $-\gamma$ | 0 | 0 | 0 |
| $R_I$ | 0 | 0 | 0 | $(1-\delta)\gamma$ | 0 | 0 | 0 | 0 | 0 |
| $R_A$ | 0 | 0 | 0 | 0 | 0 | $\gamma$ | 0 | 0 | 0 |
| $D$ | 0 | 0 | 0 | $\delta\gamma$ | 0 | 0 | 0 | 0 | 0 |

Let $\mathbf{T}$ be the matrix whose entries are given in this table, and let $t_{ij}$ denote the $i^{\text{th}}$ row $j^{\text{th}}$ column entry of $\mathbf{T}$. The diagonal entries of $\mathbf{T}$ are the rates of exit from each stage, and $t_{ij}$ with $i \neq j$ is the rate of transition from stage $j$ to stage $i$. If we let $p_i(t)$ denote the probability a given individual is in stage $i$ at time $t$ then $p(t)$ satisfies the differential equations $\frac{dp}{dt} = \mathbf{T}p$, whose solutions are the matrix exponential $e^{\mathbf{T}t}$. Specifically, entries of $e^{\mathbf{T}t}$ are the probabilities an individual is in the corresponding row-stage at time $t$ conditional on having started in the corresponding column-stage at time 0. Figure 6 gives a graphical representation of $e^{\mathbf{T}t}$ with parameters specified in Table 1 and $t$ set to 7 days and 14 days respectively. Specifically, the entries of $e^{7\mathbf{T}}$ are as follows:

| | $L_1$ | $L_2$ | $I_1$ | $I_2$ | $A_1$ | $A_2$ | $R_I$ | $R_A$ | $D$ |
|---|---|---|---|---|---|---|---|---|---|
| $L_1$ | 0.086 | 0 | 0 | 0 | 0 | 0 | 0 | 0 | 0 |
| $L_2$ | 0.211 | 0.086 | 0 | 0 | 0 | 0 | 0 | 0 | 0 |
| $I_1$ | 0.304 | 0.299 | 0.247 | 0 | 0 | 0 | 0 | 0 | 0 |
| $I_2$ | 0.168 | 0.246 | 0.345 | 0.247 | 0 | 0 | 0 | 0 | 0 |
| $A_1$ | 0.076 | 0.075 | 0 | 0 | 0.247 | 0 | 0 | 0 | 0 |
| $A_2$ | 0.042 | 0.061 | 0 | 0 | 0.345 | 0.247 | 0 | 0 | 0 |
| $R_I$ | 0.088 | 0.182 | 0.399 | 0.736 | 0 | 0 | 0.978 | 0 | 0 |
| $R_A$ | 0.023 | 0.047 | 0 | 0 | 0.408 | 0.753 | 0 | 1 | 0 |
| $D$ | 0.002 | 0.004 | 0.009 | 0.017 | 0 | 0 | 0.022 | 0 | 1 |

An individual in stage $L_2$ at time 0 is distributed at time $t = 7$ according to column 2 of the table.

If an infected individual with state distributed according to $p_X$ arrives in a community at time 0 and is placed in quarantine for a time $t_q$, then that individual's state at time $t_q$ has distribution $e^{\mathbf{T}t_q}p_X$. Further, the probability the individual is in one of the states in $\{L_1, L_2, A_1, A_2\}$ at time $t$ is given by $ue^{\mathbf{T}t}p_X$ where $u$ is the characteristic vector for undetected infections, $u = (1,1,0,0,1,1,0,0,0)$ and a range of times $t_q$. If, after $t_q$ days, the individual is still in an unobservable state $\mathcal{U}$, then quarantine has failed. Otherwise, quarantine is considered a success. Recall that, in the model, $I$ individuals have been detected by the authority, explaining why $I_1$ and $I_2$ individuals are considered as a success of the quarantine. We define the efficacy of quarantine, $c$, as the probability,

$$c = 1 - ue^{\mathbf{T}t_q}p_X, \tag{ES.8}$$

that the imported case is in either an observable state, $I_1$, $I_2$, $R_I$, or $D$, or recovered in state $R_A$. Figure 7 represents $c$ for different values of $t_q$, where $ue^{\mathbf{T}t_q}p_X$ is computed for $p_X = (0.25, 0.25, 0, 0, 0.25, 0.25, 0, 0, 0)^T$.

Furthermore, the quarantine-regulated importation rate $\lambda_q$ is expressed by

$$\lambda_q = (1 - c)\lambda. \tag{ES.9}$$
